# Health Economic Studies of Colorectal Cancer and the Contribution of Administrative Data: a Systematic Review

**DOI:** 10.1101/2020.09.29.20203679

**Authors:** E Lemmon, C Hanna, P Hall, E Morris

## Abstract

**Introduction:** Several forces are contributing to an increase in the number of people living with and surviving colorectal cancer (CRC). However, due to the lack of available data, little is known about those implications. In recent years, the use of administrative records to inform research has been increasing. The aim of this paper is to investigate the potential contribution that administrative data could have on the health economic research of CRC.

**Methods:** To achieve this aim we conducted a systematic review of the health economic CRC literature published in the United Kingdom and Europe within the last decade (2009-2019).

**Results:** Thirty-seven relevant studies were identified and divided into economic evaluations, cost of illness studies and cost consequence analyses.

**Conclusions:** The use of administrative data, including cancer registry, screening and hospital records, within the health economic research of CRC is commonplace. However, we found that this data often comes from regional databases, which reduces the generalisability of results. Further, administrative data appear less able to contribute towards understanding the wider and indirect costs associated with the disease. We have identified several ways in which various sources of administrative data could enhance future research in this area.

## 1. INTRODUCTION

Colorectal cancer (CRC) is the third most common type of cancer globally, with over 1.8 million cases diagnosed in 2018. Incidence of CRC increases with age and peak rates of new diagnoses occur in those aged 85-89. Furthermore, CRC accounts for the second highest number of deaths due to cancer. (CRUK, 2014)

Over the last forty years, technological advancements have enhanced detection and treatment for CRC, leading to improvements in survival and mortality rates (CRUK, 2014). In conjunction with the ageing population, the number of people living with and surviving cancer is expected to increase. In this way, CRC is increasingly considered a chronic condition, requiring care that stems beyond the initial phases of diagnosis and treatment.

The economic impact of better diagnosis, increasing lines of available treatment and improved survival is far reaching. Not only are there direct monetary implications for health and care service providers in terms of detection, treatment and follow up care of CRC. But there are indirect implications for patients, their families and wider society in terms of the impact of CRC on labour force participation and on both physical and mental well-being. It is crucial that we can measure these implications in order to assess the impact of CRC and to help inform policy makers decisions on how best to allocate a finite health budget.

The current availability of data to inform this understanding is somewhat limited and more often than not, data from clinical trials is used to make assumptions about the possible impact of an intervention on the entire population and ultimately inform decisions about resource allocation. Unfortunately, the generalisability of efficacy and cost-effectiveness measures from clinical trials to real life populations can be limited by sample selection, size and attrition. Furthermore, clinical trials are expensive to implement and run, and often have short follow up periods, meaning that longer term outcomes cannot be observed.

One potential solution to the issue of the generalisability of trial data to whole populations lies in the use of administrative data. That is, data that are collected routinely “by government departments and other organisations for the purposes of registration, transaction and record keeping, usually during the delivery of a service” (Woollard, 2014). Examples include hospital admissions data, education records, tax records etc. The routine collection of administrative data presents an exciting opportunity to conduct population level research that offers insights into health care resource use, costs and outcomes across a variety of domains such as education, income and retirement, through the linkage of these records to other data sets (Einav and Levin, 2014; Card et al., 2010). Moreover, administrative data can overcome the short follow up period inherent in trials by tracking individuals over time, for example as they move in and out of hospital, into long term care and even up to the end of their lives.

Despite these advantages, since administrative data are not generated for research purposes, they often lack the usual auxiliary measures that are used in social research to draw causal inference from a data set (Connelly et al., 2016). Thus, one of the central prospects for administrative data is for its use as a complementary source of information alongside clinical trials and survey data. The benefits of linking administrative records to observational data are documented elsewhere (Doiron et al., 2013).

Over the years, the potential of administrative data in research has been recognised worldwide and efforts have been made to harness that potential (Card et al., 2010; Einav and Levin, 2014). In the Nordic countries in particular, robust data sharing infrastructures have been developed to facilitate researchers in making use of administrative data sets (Connelly et al., 2016).

Moreover, the linkage aspect of administrative data has led to large data repositories emerging, where data sets are linked together and researchers can apply to access specific data sets and cohorts, to carry out their analysis (Doiron et al., 2013). Further, data repositories enhance research transparency because their indefinite storage allows for the replication of results. The success of such repositories has been made clear, for example the Western Australia Data Linkage System (WADLS) repository includes over 30 population-based datasets and has produced over 250 journal publications (Doiron et al., 2013).

Of course, the creation of such repositories is not without its challenges. In particular, any research project that uses personal health data where informed consent is not obtained from patients, may pose a risk to individual privacy. Therefore, central to the creation of a research repository is striking the appropriate balance between public benefit and patient privacy. That means being clear and transparent about the purposes of the research and its potential to generate patient or public benefit, at the same time taking measures to minimise the risk to patient privacy for example through the pseudonymisation of data.

We have identified that Scotland is in a unique position to demonstrate the potential contribution of administrative data, as well as an administrative data repository, within the health economic research of CRC. This is primarily due to the current data sharing and linkage infrastructure. Specifically, all Scottish residents have a unique Community Heath Index (CHI) number that permits the linkage of their administrative health records to one another and to other data sets.

The overarching aim of this paper is to investigate the potential contribution that administrative data could have on health economic research of CRC. To achieve this aim, the objectives were:

1. To summarise the existing health economic research of CRC in the UK and Europe;
2. To identify the types of administrative data used within this research;
3. To explore the benefits and limitations of using administrative data in this research;
4. To discuss the ways in which administrative data, using Scotland as an exemplar, could contribute to this research in the future.

In what follows we outline the methods employed for the systematic review. Section 3 presents the results and Section 4 discusses the findings and concludes.

## 2. METHODS

### Search strategy

We conducted a systematic literature search of Ovid MEDLINE® for English language articles published between 2009 and 2019. Specifically, the search strategy was as follows: 1.(Col-orectal cancer OR bowel cancer).title AND economic.abstract and cost.abstract. 2. Limit 1 to (English language AND year =“2009-2019”).

A Google Scholar search was also conducted to capture other relevant articles. We followed the PRISMA guidelines where applicable for conducting this review (PRISMA, 2020).

### Selection criteria

Full text publications of health economic studies were included when available in English language. The definitions of health economic studies are outlined in Table 1. Articles that were not carried out in the Europe or the UK were excluded. Further, review articles were also excluded.

**Table 1:** Definition of health economic studies included in final review

| **Study** |  |  | **Description** |
| --- | --- | --- | --- |
| **Budget Impact Analysis (BIA)** |  |  | Budget impact analyses assess the affordability of a novel health care intervention or policy change applied to a specific healthcare budget, at an aggregate population level. |
| **Cost Comparison (Cost Minimisation) (CC)** |  |  | CC is a method of comparing the costs of two or more interventions when the health outcomes of the interventions are assumed to be the same. |
| **Cost of Illness (Burden of Illness) (COI)** |  |  | COI studies attempt to quantify the costs of a specific disease. This might be for the entire disease pathway or for parts of it. Unlike EEs, they do not attempt to compare costs for competing interventions rather, they provide an estimate of the cost given the existing provision of care. |
| **Economic Evaluation** |  |  | Economic evaluation aims to calculate the costs and benefits of an intervention or treatment, in order to establish whether it is cost effective and thus inform investment in services. There are four main types of economic evaluation, which differ in terms of how they measure outcomes: |
| Cost Benefit Analysis (CBA) |  |  | In CBA, health outcomes are measured in monetary units. |
| Cost Effectiveness Analysis (CEA) |  |  | In CEA, outcomes are measured in natural or health units such as life years gained or cancers detected. |
| Cost Utility Analysis (CUA) |  |  | CUA is a special type of CEA in which outcomes are measured in preference based health outcomes such as Quality Adjusted Life Years (QALY). |
| Source:Adapted from York Health Economics Consortium Glossary (York Health Economics Consortium, 2016) |  |  |  |

### Data extraction

The articles were grouped into the study groups as outlined in Table 1. A proforma was used to extract the relevant data from each article within these groups. For all types of studies, the country, perspective taken, method employed, data sources used (including administrative data), types of costs included, costs data sources used and the part of the CRC pathway under study were extracted. For the EEs, the type of evaluation was also noted.

## 3. RESULTS

### Literature search results

Fig. 1 below outlines the PRISMA (PRISMA, 2020). flow diagram of the search strategy results.

**Figure 1:**
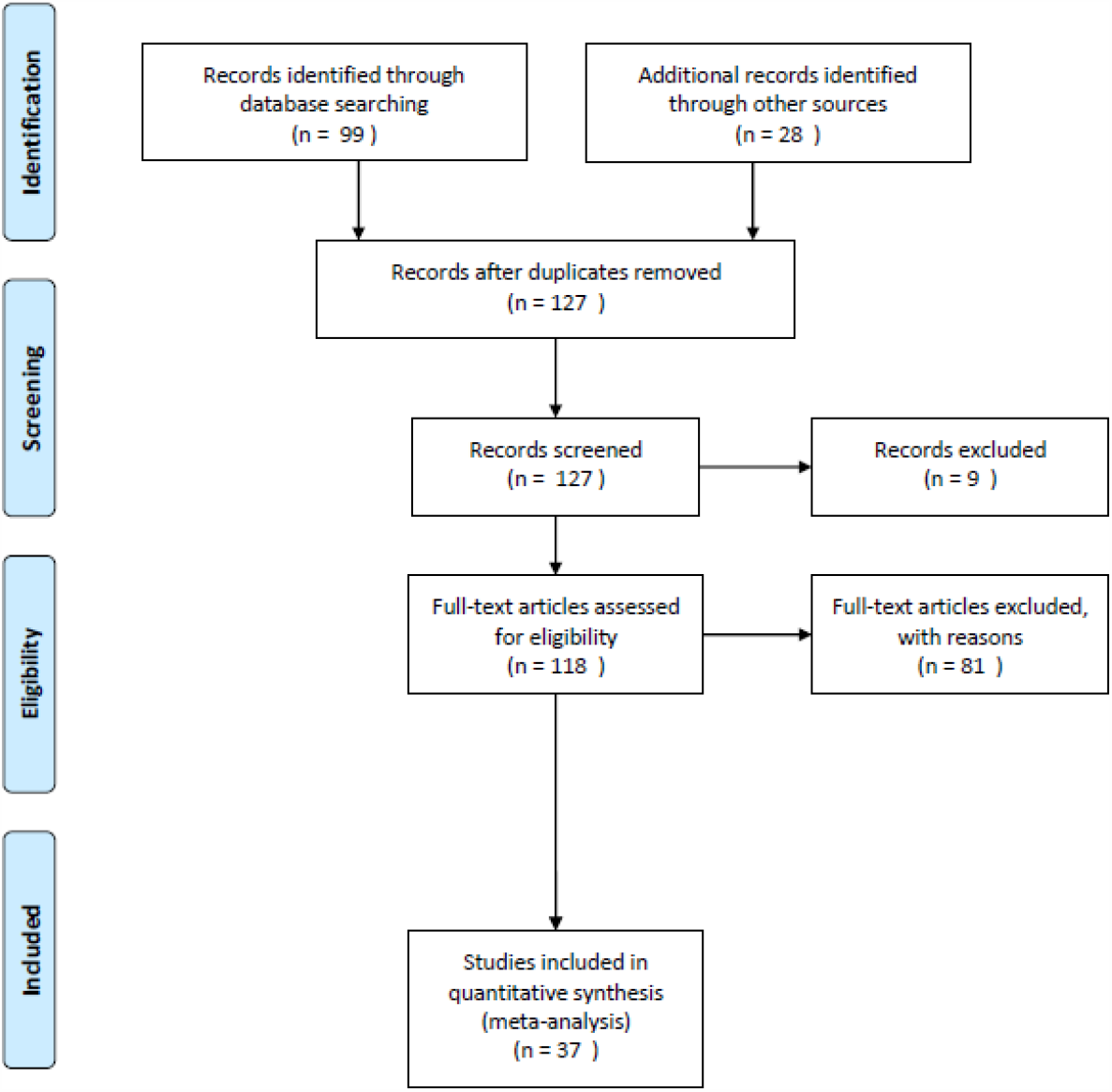
Literature Search Results.

The search identified a total of 127 potentially relevant articles to be screened. After removing duplicates and non applicable applications (i.e. non-economic studies), 118 articles remained. Of those, 25 were review articles and 56 were non EU/UK, leaving a final 37 studies that matched our inclusion criteria.

The articles were almost equally split between EEs (n = 19) and costing studies (n = 18). As per Table 1 the costing studies were categorised into Cost of Illness (COI) studies (n = 13) and Cost Comparison (CC) studies (n= 5).

### Summary of the existing health economic research of CRC in the UK and Europe

Table 2, Table 3 and Table 3 present the EEs, COI and CCs included in the final review. Overall, 51% of the total studies included were EEs, 35% were COI studies and 14% were CC studies.

**Table 2:** UK/EU Economic Evaluations, n = 19

| Reference | Country | Perspective | Evaluation | Method | Data Sources | Admin Data | Costs | Data Sources: Costs | Pathway |
| --- | --- | --- | --- | --- | --- | --- | --- | --- | --- |
| Arrospeide et al. (2018) | Basque | Health care system | CUA, BIA | Semi-Markov Microsimulation | Screening data, cancer registry, national statistics | Yes | Direct | Accounting system | Screening- end of life |
| Atkin et al. (2017) | UK | Health and social care system | CUA | Patient Level Simulation | Hospital records, previous studies, life tables | Yes | Direct | National tariffs | Surveillance- end of life |
| Asseburg et al. (2011) | Germany | Health insurance perspective | CEA | Patient Level Simulation | Previous studies, expert opinion | Yes (costs only) | Direct | National tariffs, market prices | Treatment - 10 years after |
| Bullement et al. (2018) | England/Wales | Health care system | CEA, CUA | Partitioned Survival Model | Previous studies | Yes (costs only) | Direct | Market prices (MIMS), national tariffs, previous study | Treatment- end of life |
| Halligan et al. (2015) | England | Health care system-secondary care | CEA | Descriptive | Primary data collection | Yes (costs only) | Direct | National tariffs, BNF | Diagnosis-5 years after |
| Kearns et al. (2014) | England | Health and social care system | CEA, CUA | Markov Model | Previous studies, screening data | Yes | Direct | Previous study | Screening- end of life |
| Lansdorp-Vogelaar et al. (2018) | Netherlands | Societal perspective | CEA | Semi-Markov Microsimulation | Previous studies, life tables | No | Direct & indirect | Previous studies | Screening- end of life |
| Matter-Walstra et al. (2016) | Switzerland | Health care system | CEA | Descriptive, regression | Previous study | No | Direct | Primary data collection | Treatment (and overall survival) |
| Michalopoulos et al. (2013) | Greece | Not stated | CUA | Descriptive, regression | Primary data collection | No | Direct | Primary data collection | Surgery |
| Murphy et al. (2017) | England | Health care system | CUA, BIA | Markov Model | Previous studies, life tables | Yes (costs only) | Direct | National tariffs, screening data, previous study | Screening-end of life |
| Pil et al. (2016) | Belgium | Societal perspective | CUA, BIA | Decision Tree & Markov Model | Previous studies, cancer registry, screening data, national statistics | Yes | Direct & indirect | Official Belgian costs of medical procedures, previous studies | Screening - 50 years after |
| Pilgrim et al. (2009) | England | Not mentioned | CUA | Discrete Event Simulation | Previous studies, hospital records, expert opinion | Yes | Direct | National tariffs, previous studies | Screening- end of life |
| Rao et al. (2018) | UK | Health care system | CUA | Decision Tree & Markov Model | Previous studies, hospital records, life tables | Yes | Direct | National tariffs | Treatment- end of life |
| Rautenberg et al. (2014) | Germany | Health insurance perspective | CEA | Descriptive | Previous studies | No | Direct | Market prices | Treatment |
| Robles-Zurita et al. (2018) | UK<sup>1</sup> | Health and social care system | CUA | Partitioned Survival Model | Previous study | Yes (costs only) | Direct | National tariffs, primary collection | Treatment- 8 years after |
| Sobhani et al. (2011) | France | Health care payer | CUA | Markov Model | Previous studies, national statistics | No | Direct | Previous studies | Screening - end of life |
| Tilson et al. (2012) | Ireland | Health care payer | CEA | Decision Tree Model | Previous studies, cancer registry, hospital records, expert opinion | Yes | Direct | National tariffs, previous studies, hospital finance department | Diagnosis-5 years after |
| van der Meulen et al. (2018) | Netherlands | Third party payer | CUA | Semi-Markov Microsimulation | Previous studies, life tables | No | Direct | Previous study | Screening- end of life |
| Whyte et al. (2012) | England | Health care system | CUA | State Transition Model | Previous studies, screening data, expert opinion | Yes | Direct | National tariffs, previous study | Screening- end of life |

**Table 3:** Cost of Illness, n = 13

| Reference | Country | Perspective | Method | Data Sources | Admin Data | Costs | Data Sources: Costs | Pathway |
| --- | --- | --- | --- | --- | --- | --- | --- | --- |
| Bending et al. (2010) | England | Not mentioned | Service pathway model | Hospital records, previous studies | Yes | Direct | National tariffs | Diagnosis- end of life |
| Corral et al. (2016) | Spain | Not mentioned | Retrospective cohort, survival analysis | Cancer registry, clinical admin system | Yes | Direct | Hospital Unit Costs | Diagnosis- end of life |
| Francisci et al. (2013) | Italy | Not mentioned | Retrospective cohort | Cancer registry, hospital data | Yes | Direct | National tariffs | Diagnosis- end of life |
| Giuliani et al. (2012) | Italy | Not mentioned | Retrospective cohort | Clinical records, medical histories | Yes | Direct | Market prices | Treatment (metastatic) |
| Hall et al. (2015) | England | Not mentioned | Retrospective cohort, regression analysis | Cancer registry, PROMs, hospital records | Yes | Direct | Hospital finance data |  |
| Hanly et al. (2013) | Ireland | Societal | Descriptive | Primary data | Yes | Indirect | National wages | 12-36 months post diagnosis |
| Jean-Claude et al. (2012) | France | Not mentioned | Retrospective cohort, BIA | Previous study | No | Direct | Previous study | Surgery |
| Laudicella et al. (2016) | England | Not mentioned | Retrospective cohort | Hospital records, cancer registry | Yes | Direct | Hospital Unit Costs | Hospital care |
| Lejeune et al. (2009) | France | Health system (France) | Retrospective cohort | Cancer registry, physician records | Yes | Direct & indirect | Reimbursement Prices | Surgery- 3 years after |
| Macafee and Whynes (2009) | England | Treatment level | Retrospective cohort | Hospital records | Yes | Direct | National tariffs, hospital finances, pharmacy | Hospital care |
| Mar et al. (2017) | Spain | Health care system | Retrospective cohort, survival analysis | Hospital records | Yes | Direct | National tariffs o | Treatment (metastatic & non-metastatic)- end of life |
| ÓCéilleachair et al. (2012) | Ireland | Patient | Qualitative interviews | Primary data | Yes (to identify participants) | NA | NA | Within 12 months of diagnosis |
| ÓCéilleachair et al. (2017) | Ireland | Patient | Retrospective cohort | Cancer registry, primary data | Yes | Indirect | Primary collection | Diagnosis - initial follow up |

Around 47% (n = 9) of the EEs were conducted in the UK and the majority (68%) of EEs conducted a CUA, whilst the remainder chose to implement a CEA. As is standard within an EE framework, the majority of studies accompany their EE with a decision tree, Markov model or simulation model, in order to extrapolate CRC costs and outcomes over time, for example up until the end of life.

In EE, it is standard practice to explicitly state the perspective of the evaluation. Of the 19 EE’s identified, 53% (n = 10) take a health care system perspective. Other perspectives taken include a health care payer perspective, health insurance perspective or societal perspective. The perspective taken influences the types of costs that are included. As a result, the vast majority (89%) of studies only include direct costs associated with the delivery of care. The two studies which take a societal perspective, Lansdorp-Vogelaar et al. (2018) and Pil et al. (2016), also incorporate indirect costs i.e additional costs encountered by the patients such as loss of earnings. In terms of which part of the CRC pathway is investigated, the most common evaluations are conducted on screening programmes. In particular, 45% (n = 9) of the included studies evaluate the cost-effectiveness of different CRC screening programmes. A further 40% (n = 7) look at the cost-effectiveness of treatment for CRC, including curative treatment and treatment for metastatic disease. A smaller proportion of the EE’s, 10% (n = 2), look at diagnosis and 5% (n = 1) at surveillance of adenomas. As most studies use Markov and Microsimulation models, they tend to model outcomes and costs beyond the initial pathway starting point, either until the end of life or an alternative long term end point e.g. 50 year follow up. In addition to conducting a CUA to assess the value of a health care intervention, three of the EEs conducted a budget impact analysis (BIA) to assess the affordability of the intervention for a specific health care budget (Arrospide et al., 2018; Murphy et al., 2017; Pil et al., 2016).

In contrast to the evaluations, only 31% (n = 4) of the COI articles identified were UK based (England only). Ireland accounted for almost a quarter of the studies (n = 3), followed by Italy (n = 2), France (n = 2) and Spain (n = 2). The majority of COI articles conducted retrospective cohort analyses. This involves looking at historical data to identify a cohort of patients, for example those with metastatic CRC, and costing their use of health care resources.

In addition, COI studies were less likely to mention which perspective the analysis is conducted from. However, like EEs, the COI studies tended to focus on direct costs. Only two COI papers looked solely at indirect costs (Hanly et al., 2013; ÓCéilleachair et al., 2017) and one included both direct and indirect costs (Lejeune et al., 2009). With respect to the particular part of the CRC care pathway costed, there was considerable variation within the COI studies. Three looked at the pathway from diagnosis to end of life (Bending et al., 2010; Corral et al., 2016; Francisci et al., 2013). Others focused on diagnosis, but only looked at costs from diagnosis up to a pre-specified time point e.g. 12 months post diagnosis or within 12 months of initial diagnosis. Two studies looked at all hospital care throughout the care pathway (Laudi- cella et al., 2016; Macafee and Whynes, 2009). One study focussed on treatment of metastatic and non-metastatic disease up until the end of life Mar et al. (2017) whilst another focussed on costs of treating metastatic disease alone (Giuliani et al., 2012). Similarly, one study looked at the cost of surgery alone (Jean-Claude et al., 2012)and another at the costs from surgery up to three years post-surgery (Lejeune et al., 2009). One COI study also conducted a BIA for patients who underwent CRC surgery in French non-profit hospitals (Jean-Claude et al., 2012).

Of the CC studies identified in Table 4, two of the CC studies are from Italy and the remaining three are from Greece, Sweden and Germany. As with the COI studies, the predominant methodology applied in the CCs is a retrospective cohort approach. In terms of the perspective, the majority of the CC studies conduct their analyses from the perspective of the health system in which they are based. One of the studies takes the perspective of the health care payer and another takes both a health policy maker and a societal perspective. Once again, the focus on costs is mainly on direct costs, however two CC papers also incorporate indirect costs. The majority of CC studies focus on the treatment part of the CRC pathway. One paper focuses on surgery and another on screening up until the end of life.

**Table 4:** Cost Comparisons, n = 5

| Reference | Country | Perspective | Method | Data Sources | Admin Data | Costs | Data Sources: Costs | Pathway |
| --- | --- | --- | --- | --- | --- | --- | --- | --- |
| Berto et al. (2012) | Italy | Health care system (Italy) | Retrospective cohort | Italian reference centres | Yes | Direct | National tariffs, hospital accounting system, market prices | Surgery |
| De Portu et al. (2010) | Italy | Health care system (Italy) | Retrospective cohort | Primary collection | No | Direct | National tariffs, market prices | Treatment (metastatic) |
| Maniadakis et al. (2009) | Greece | Health care system (Greece) | Descriptive, survival analysis | Previous RCT | Yes (costs only) | Direct & indirect | Hospital accounting system, reimbursement prices | Treatment (curative) |
| Pettersson et al. (2012) | Sweden | Health care payer (Gothemberg) | Retrospective cohort | Hospital data | Yes | Direct | Local hospital costs, pharmacy costs | Treatment (metastatic) |
| Tscheulin and Dreves (2010) | Germany | Health policy maker & societal | Step-wise cost comparison | Population data | Yes (costs only) | Direct & indirect | Previous studies, physician fee schedule | Screening- end of life |

### Types of administrative data used within the health economic research of CRC in the UK and Europe

The papers identified use a mixture of data sources including administrative data, national statistics, other randomised control trials (RCTs), previous literature, expert opinion, and in some cases primary data collection. Table 5 below outlines the administrative data sources that appeared most frequently in the studies.

**Table 5:** Administrative data sources

| **Data source** | **Description** |
| --- | --- |
| Cancer registry data | Cancer registries contain a record of all cases of new cancer diagnoses in one centralised system. They tend to include information on cancer diagnoses and treatment, allowing a country to monitor cancer incidence and survival, and any emerging trends, over a long period of time. Registries also include patient level demographics, permitting analyses of diagnoses by age, gender and stage distribution. They can also include information on cancer related mortality. |
| Screening programme data | Screening programme datasets provide a wealth of information including participation and compliance rates, adenoma and CRC detection rates, specificity and sensitivity, as well as information on surveillance. In some cases, follow up data are also available, for example on colonoscopies and flexible sigmoidoscopy. Follow up data provide information on participation, detection and complications. |
| Routine hospital records | Routine hospital records provide information on any acute hospital admission experienced by a patient, including length of stay and procedure codes. Moreover, hospital records often include additional information about an individuals primary and secondary diagnoses, allowing the researcher to gather more information about patient co-morbidity and other procedures and medications related to or unrelated to their cancer diagnosis. |
| Costs databases | Administrative costs data are collected in the form of national tariffs for the reimbursement of the provision of hospital services and in hospital accounting systems. These systems are usually updated annually and therefore provide robust and up to date estimates of unit costs for economic analyses. |

Within the EEs, 68% (n = 13) utilise administrative data. In general, the administrative data are used to inform particular parameters in the Decision Trees, Markov and simulation models. For example, cancer registry data are used to inform incidence and prevalence parameters. Of the EEs which use administrative data, five evaluate screening programmes and use administrative screening programme data in their analysis. This clearly reflects the effort in many European countries in recent years to detect cancer as early as possible for those at the highest risk by rolling out national screening programmes for CRC. As a result, a multitude of administrative screening datasets have been created and researchers have capitalised on this opportunity.

It is also common for those studies to combine the screening data with other administrative datasets. In particular, Arrospide et al. (2018) combine data from the Basque screening programme with cancer registry data to evaluate the Basque CRC Screening Programme. Pil et al. (2016) use data from the Belgian Government Screening Programme alongside the Belgian Cancer Registry in their analysis of a population based CRC screening programme. Finally, in their evaluation of different surveillance strategies for patients with intermediate-grade adenomas, Atkin et al. (2017) use routine hospital records together with data from the English Bowel Cancer Screening Pilot.

At the same time, the extent to which administrative data are used within the EEs varies considerably and no one study relies exclusively on routine data. For example, Atkin et al. (2017) use routine hospital records linked to cancer registry data to inform many of the parameters in their patient level simulation model, whilst Rao et al. (2018) use routine hospital records solely for the purposes of informing their parameter on postoperative mortality. Furthermore, several EEs use administrative costs data only. In every EE, the existing literature or previous RCTs are also used to inform specific model parameters.

The COI studies utilise administrative data more often compared to the EEs. In particular, 11 of the 12 COI papers identified in Table 3 use administrative data. In comparison to the EE’s, where much of the administrative data comes from screening programmes, the main sources of administrative data in the COI studies come from cancer registries and routine hospital records. In the majority of cases, the COI studies use cancer registry data to identify a cohort of patients to be analysed. The registry data provide information on CRC diagnosis, staging, location of tumour, date of diagnosis etc. This information is then linked with routine hospital records that contain information on treatment, co morbidities, complications and recurrence etc.

In addition to using cancer registries and clinical information systems to analyse CRC cohorts, several COI studies use registry data as a means of identifying patients to invite them to participate in a survey or interview. Specifically, Hanly et al. (2013) and ÓCéilleachair et al. (2017) use Irish cancer registry data to identify individuals with primary, invasive CRC in order to invite them to complete postal questionnaires. Survey responses are then combined with clinical information from the cancer registry to conduct statistical analysis. Similarly, ÓCéilleachair et al. (2012) used hospital records from six participating sites to identify patients who would be eligible to take part in interviews for their qualitative analysis of the inter-relationships between the economic and emotional consequences of CRC. Overall, compared to EEs, COI studies are more likely to rely exclusively on administrative data (Corral et al., 2016; Francisci et al., 2013; Giuliani et al., 2012; Laudicella et al., 2016; Lejeune et al., 2009; Macafee and Whynes, 2009; Mar et al., 2017). Finally, almost all of the CC studies use administrative data of some sort. As with the EEs, some use administrative data in the form of costs databases only and like the COI studies some use administrative hospital records.

## 4. DISCUSSION AND CONCLUSION

### The benefits and limitations of using administrative data within the health economic research of CRC in the UK and Europe

### Benefits

Clearly, one area in which administrative data have been particularly powerful is in evidence on the cost effectiveness of various screening strategies for CRC, which has resulted from the evolution of national screening programmes throughout Europe. Data from these programmes has been used to inform and update many of the crucial parameters used in the models that accompany EEs of screening programmes. This evidence base invariably demonstrates the feasibility and potential of collecting administrative data on this scale to inform other parts of the treatment pathway for CRC.

At the same time, administrative cancer registry data have proved to be useful in terms of defining and identifying cohorts for costing studies and again for informing vital parameters such as disease prevalence, treatment and outcomes. Many EE’s have also taken advantage of the power of data linkage by linking administrative records to data form participants in RCTs.

Furthermore, since providing estimates of costs is central to conducting both EE’s and costing analyses, the emergence of costs databases have proved to be a valuable source of information on costs for all areas of economic research into CRC. Specifically, 43% (n=16) of the studies identified used administrative costs databases.

In the health economics literature, costs tend to be divided into two categories. Those are, direct costs and indirect costs. Direct costs are those which relate directly to patient care such as a hospital stay, whilst indirect costs occur outside the delivery of patient care, such as lost productivity or foregone wages.

The administrative costs databases have proved particularly powerful in the studies that include direct costs. In particular, the costing approaches implemented in those papers are consistent with the existence of European Disease Related Group (DRG) type systems for reimbursing hospitals for their services. Therefore, unsurprisingly, many of them implement a ‘top-down’ costing approach by using national tariffs based on DRGs to attach monetary values to patients resource utilisation (Špacírová et al., 2020). This highlights the potential for administrative data to contribute to understanding the costs of delivering CRC care.

At the same time, one study used administrative data to inform the calculation of indirect costs. Specifically, Lejeune et al. (2009) use hospital records data to measure the distance travelled to and from the patients home to consult with their GP or gastroenterologist, which was then used to calculate indirect costs to the patient.

Finally, the merit of using administrative data for the purposes of BIA is clear. In the three EEs (Arrospide et al., 2018; Pil et al., 2016; Murphy et al., 2017) and one COI (Jean-Claude et al., 2012) who undertook BIAs, location specific estimates of population size, age-specific disease incidence, resource use and location-specific costs were acquired from various administrative data sources to permit analyses that were relevant and useful to the budget holder in question. In an era of increasing austerity and budget cuts, using administrative data within BIAs to more accurately predict the affordability of introducing novel interventions into a fixed budget healthcare system will ensure more efficient allocation of resources. Using locally or nationally collected administrative data for the purposes of BIA is particularly useful because this will make any analysis more relevant and useful to the budget holder in question.

### Limitations

Having said that, we have identified some areas where the use of administrative data has been limited. For example, although one of the main advantages of using routine records in research is their ability to capture large populations over long periods of time, we find little evidence that this is the case for the health economics literature on CRC. Specifically, only one costing study used routine records to capture an entire population over a long period of time (Laudicella et al., 2016). Excluding this example, the maximum sample size identified is less than a few thousand and in most cases, the populations under study come from a single hospital or administrative area. At the same time, many of the costing studies identified look at one specific part of the disease pathway with a limited follow up period. Overall, it appears that the power of administrative data to provide evidence for whole populations, spanning the entire disease pathway and follow up for survivors, is yet to be harnessed.

Related to this, we found a lack of evidence on the wider costs associated with CRC, particularly with respect to social care and indirect costs such as unpaid care. For example, although evidence shows that many cancer patients need social care as a direct consequence of their condition and the consequences of its treatment, none of the papers identified look at the use of social care services by CRC patients (MacMillan Cancer Support, 2015).

Furthermore, few papers explored indirect costs. In particular, only two EEs explicitly take a societal perspective and therefore include both direct and indirect costs of care (Lansdorp- Vogelaar et al., 2018; Pil et al., 2016). Within the COI studies, Hanly et al. (2013) and ÓCéil- leachair et al. (2017) focus exclusively on indirect costs, whilst Lejeune et al. (2009) include direct and indirect costs. Further, the CC’s carried out by Maniadakis et al Maniadakis et al. (2009) and Tscheulin and Drevs (2010), also include both direct and indirect costs. The lack of inclusion of indirect costs overall is not surprising given that they are notoriously difficult to measure. However, of those who did, the use of administrative data was even less likely. Clearly, measuring indirect costs is challenging in itself but in addition to this, the administrative data appear less able to contribute to studies which include the indirect costs of CRC. This highlights a key limitation of administrative records in their ability to capture indirect costs.

Finally, it appears that administrative data are less able to contribute when it comes to measuring patient health related quality of life (HRQoL) and preferences for those health states, which is vital particularly in EEs. In most cases, studies look to previous literature, often going back several years, for this information. As administrative data are not collected for the purposes of research, it isn’t surprising that they lack the types of measures needed to capture patient outcomes in the way that is needed for EEs. Nonetheless, recent developments in tools to capture Patient Reported Outcome Measures (PROMS) on symptoms, condition and quality of life might be used to measure outcomes in CRC patients.

In summary, it is not uncommon for the health economic research of CRC to utilise administrative records to aid EEs and costing analyses, and undoubtedly, they can offer a wealth of information about an individuals CRC diagnoses, subsequent treatment and follow up over time. However, there appear to be several limitations to their use and gaps in the existing evidence. The following section considers how Scottish administrative data might mitigate those limitations and fill in some of the gaps in the evidence.

### The contribution of Scottish administrative data within the health economic research of CRC in the UK and Europe

As discussed in Section 1, Scotland is in a prime position to demonstrate the contribution of administrative data, in particular due to its data sharing and data linkage infrastructure. In theory, this infrastructure means that all health data sets can easily be linked to one another and to administrative data sets in other domains, for example social care.

In October 2018, the Public Benefit Privacy Panel for Health and Social Care (PBPP) approved a project to link several administrative data sets for CRC patients in Scotland (Study number:1718-0026), in order to conduct research into the economics of CRC. This project is part of a wider Cancer Research UK funded project, Bowel Cancer Intelligence UK, which has been granted permission from the Research Ethics Service of the Health Research Authority for a COloRECTal Repository (CORECT-R) (BCIUK, 2018). The Scottish CORECT-R data provides a useful platform on which to demonstrate the possible contribution of administrative data and a CRC repository to the health economic research of CRC within the UK and Europe.

First and foremost, Scottish administrative data could contribute to the evidence by simply providing evidence for Scotland. Despite the wealth of administrative data sets and data infrastructure in existence in Scotland, there appears to be limited health economics research of CRC within the Scottish context. In particular, only two studies identified in this review used Scottish administrative records (Atkin et al., 2017; Robles-Zurita et al., 2018). In both studies, Scotland was represented alongside data from other countries. Thus, a Scottish CRC data repository would afford the opportunity for health economic research into CRC in Scotland to be realised.

Secondly, the use of Scottish administrative data could be used to inform and update common model parameters used in both EEs and costing studies, using data that reflects current practice for an entire population. For example, prevalence and incidence rates, durations of treatments, survival outcomes etc. This information could not only be useful for other health economic studies of CRC in Scotland, but also for other nations in the UK and potentially other European countries who have similar demographics and health systems.

Thirdly, the data linkage infrastructure in Scotland would mean that all relevant health data sets can be linked to one another. In particular, cancer registry, cancer treatment, screening, outpatient and inpatient, prescriptions, accident and emergency, GP data and more. This level of information would potentially capture the patients entire CRC journey through the health care system, pre and post diagnosis, allowing for the more precise measurement of the key inputs into health economic studies of CRC. Furthermore, health data sets can also be linked to other administrative databases like social care and Department for Work and Pensions (DWP) data. These linkages to other administrative data sets outwith the health care system could provide additional information about a patients experience which could again be used to inform health economic research in this area, both within and out-with the UK.

Finally, and related to the linkage opportunities, the use of Scottish administrative data could enhance the evidence base on other direct and indirect costs related to CRC. Specifically, the linkage to social care records could be particularly powerful in this respect. As highlighted earlier, none of the studies we identified looked at the use of longer term social care services by CRC patients, despite existing evidence showing that many cancer patients need social care as a direct consequence of their condition and the consequences of its treatment (MacMillan Cancer Support, 2015). In Scotland, Local Authorities are required to routinely collect information on all social care services delivered to people within their area. This data could be used to provide evidence on other non-health related direct costs associated with CRC, again both during treatment and beyond. In addition, as part of the social care data collection, an indicator of the presence of an unpaid carer is collected for social care clients. This information could be useful for understanding the indirect costs associated with a CRC diagnosis, in terms of the reliance on unpaid carers to provide additional care and support.

## Conclusion

In conclusion, the use of administrative data is common within the UK and EU health economic research on CRC. In particular, cancer registry, screening and routine hospital records were commonly used. In the EE’s, administrative data tended to be supplemented with data from the clinical trial under study and/or from the existing literature. Costing studies were more likely to rely heavily on administrative records. Overall, we find that although administrative data are present, they do not appear to being used to their full potential and administrative data, including data repositories, within the UK and Europe could have a significant impact on research in this area. Scotland, in particular, may provide a valuable exemplar to unlock this potential.

Going forward, there are areas in which the administrative data are simply not able to fill in the gaps, especially around indirect costs and also on the well-being of patients and their caregivers. Since administrative data are collected for non-research purposes, it is not surprising that this information is lacking. One solution would be to collect this information directly from patients and their caregivers, perhaps via a survey or qualitative interviews, and this data could then be linked to the administrative data.

## Data Availability

Data sharing not applicable to this article as no datasets were generated or analysed during the current study

## References

1. Arrospide, A., Idigoras, I., Mar, J., de Koning, H., van der Meulen, M., Soto-Gordoa, M., Martinez-Llorente, J.M., Portillo, I., Arana-Arri, E., Ibarrondo, O., et al., 2018. Costeffectiveness and budget impact analyses of a colorectal cancer screening programme in a high adenoma prevalence scenario using miscan-colon microsimulation model. BMC cancer 18, 464.

2. Asseburg, C., Frank, M., Köhne, C.H., Hartmann, J.T., Griebsch, I., Mohr, A., Osowski, U., Schulten, J., Mittendorf, T., 2011. Cost-effectiveness of targeted therapy with cetuximab in patients with k-ras wild-type colorectal cancer presenting with initially unresectable metastases limited to the liver in a german setting. Clinical therapeutics 33, 482–497.

3. Atkin, W., Brenner, A., Martin, J., Wooldrage, K., Shah, U., Lucas, F., Greliak, P., Pack, K., Kralj-Hans, I., Thomson, A., et al., 2017. The clinical effectiveness of different surveillance strategies to prevent colorectal cancer in people with intermediate-grade colorectal adenomas: a retrospective cohort analysis, and psychological and economic evaluations. NIHR Journals Library.

4. BCIUK, 2018. Bowel cancer intelligence uk. https://bci.leeds.ac.uk/corect-r/ Accessed: 20.07.2020.

5. Bending, M.W., Trueman, P., Lowson, K.V., Pilgrim, H., Tappenden, P., Chilcott, J., Tappenden, J., 2010. Estimating the direct costs of bowel cancer services provided by the national health service in england. International journal of technology assessment in health care 26, 362–369.

6. Berto, P., Lopatriello, S., Aiello, A., Corcione, F., Spinoglio, G., Trapani, V., Melotti, G., 2012. Cost of laparoscopy and laparotomy in the surgical treatment of colorectal cancer. Surgical endoscopy 26, 1444–1453.

7. Bullement, A., Underhill, S., Fougeray, R., Hatswell, A.J., 2018. Cost-effectiveness of trifluridine/tipiracil for previously treated metastatic colorectal cancer in england and wales. Clinical colorectal cancer 17, e143–e151.

8. Card, D., Chetty, R., Feldstein, M.S., Saez, E., 2010. Expanding access to administrative data for research in the united states. American Economic Association, Ten Years and Beyond: Economists Answer NSF’s Call for Long-Term Research Agendas.

9. Connelly, R., Playford, C.J., Gayle, V., Dibben, C., 2016. The role of administrative data in the big data revolution in social science research. Social Science Research 59, 1–12.

10. Corral, J., Castells, X., Molins, E., Chiarello, P., Borras, J.M., Cots, F., 2016. Long-term costs of colorectal cancer treatment in spain. BMC health services research 16, 56.

11. CRUK, 2014. Cancer research uk, bowel cancer incidence statistics (2014-2016). https://edin.ac/32Abfal Accessed:20.07.2020.

12. De Portu, S., Mantovani, L., Ravaioli, A., Tamburini, E., Bollina, R., Cozzi, C., Grimaldi, A., Testa, T., Bianchessi, C., Cartenì, G., 2010. Cost analysis of capecitabine vs 5-fluorouracil-based treatment for metastatic colorectal cancer patients. Journal of Chemotherapy 22, 125–128.

13. Doiron, D., Raina, P., Fortier, I., et al., 2013. Linking canadian population health data: maximizing the potential of cohort and administrative data. Canadian journal of public health 104, e258–e261.

14. Einav, L., Levin, J., 2014. The data revolution and economic analysis. Innovation Policy and the Economy 14, 1–24.

15. Francisci, S., Guzzinati, S., Mezzetti, M., Crocetti, E., Giusti, F., Miccinesi, G., Paci, E., Angiolini, C., Gigli, A., 2013. Cost profiles of colorectal cancer patients in italy based on individual patterns of care. BMC cancer 13, 329.

16. Giuliani, J., Indelli, M., Marzola, M., Raisi, E., Frassoldati, A., 2012. The management of skin toxicity during cetuximab treatment in advanced colorectal cancer: how much does it cost? a retrospecive economic assessment from a single-center experience. Tumori Journal 98, 408–412.

17. Hall, P., Hamilton, P., Hulme, C., Meads, D., Jones, H., Newsham, A., Marti, J., Smith, A., Mason, H., Velikova, G., et al., 2015. Costs of cancer care for use in economic evaluation: a uk analysis of patient-level routine health system data. British journal of cancer 112, 948.

18. Halligan, S., Dadswell, E., Wooldrage, K., Wardle, J., von Wagner, C., Lilford, R., Yao, G.L., Zhu, S., Atkin, W., 2015. Computed tomographic colonography compared with colonoscopy or barium enema for diagnosis of colorectal cancer in older symptomatic patients: two multicentre randomised trials with economic evaluation (the siggar trials). NIHR Journals Library.

19. Hanly, P., Céilleachair, A.Ó., Skally, M., O’Leary, E., Staines, A., Kapur, K., Fitzpatrick, P., Sharp, L., 2013. Time costs associated with informal care for colorectal cancer: an investigation of the impact of alternative valuation methods. Applied health economics and health policy 11, 193–203.

20. Jean-Claude, M., Emmanuelle, P., Juliette, H., Michèle, B., Gérard, D., Eric, F., Xavier, H., Bertrand, L., Jean-Fabien, Z., Yves, P., et al., 2012. Clinical and economic impact of malnutrition per se on the postoperative course of colorectal cancer patients. Clinical nutrition 31, 896–902.

21. Kearns, B., Whyte, S., Chilcott, J., Patnick, J., 2014. Guaiac faecal occult blood test performance at initial and repeat screens in the english bowel cancer screening programme. British journal of cancer 111, 1734.

22. Lansdorp-Vogelaar, I., Goede, S.L., Bosch, L.J., Melotte, V., Carvalho, B., van Engeland, M., Meijer, G.A., de Koning, H.J., van Ballegooijen, M., 2018. Cost-effectiveness of highperformance biomarker tests vs fecal immunochemical test for noninvasive colorectal cancer screening. Clinical gastroenterology and hepatology 16, 504–512.

23. Laudicella, M., Walsh, B., Burns, E., Smith, P.C., 2016. Cost of care for cancer patients in england: evidence from population-based patient-level data. British journal of cancer 114, 1286.

24. Lejeune, C., Binquet, C., Bonnetain, F., Mahboubi, A., Abrahamowicz, M., Moreau, T., Raikou, M., Bedenne, L., Quantin, C., Bonithon-Kopp, C., 2009. Estimating the cost related to surveillance of colorectal cancer in a french population. The European Journal of Health Economics 10, 409–419.

25. Macafee, J.W.J.S., Whynes, D., 2009. Hospital costs of colorectal cancer care. Clinical Medicine: Oncology 3, 27–37.

26. MacMillan Cancer Support, 2015. Hidden at home: The social care needs of people with cancer. https://edin.ac/39HvTFg Accessed: 20.07.2020.

27. Maniadakis, N., Fragoulakis, V., Pectasides, D., Fountzilas, G., 2009. Xelox versus folfox6 as an adjuvant treatment in colorectal cancer: an economic analysis. Current medical research and opinion 25, 797–805.

28. Mar, J., Errasti, J., Soto-Gordoa, M., Mar-Barrutia, G., Martinez-Llorente, J.M., Domínguez, S., García-Albás, J.J., Arrospide, A., 2017. The cost of colorectal cancer according to the tnm stage. Cirugía Española (English Edition) 95, 89–96.

29. Matter-Walstra, K., Schwenkglenks, M., Betticher, D., von Moos, R., Dietrich, D., Baertschi, D., Koeberle, D., 2016. Bevacizumab continuation versus treatment holidays after first-line chemotherapy with bevacizumab in patients with metastatic colorectal cancer: A health economic analysis of a randomized phase 3 trial (sakk 41/06). Clinical colorectal cancer 15, 314–320.

30. van der Meulen, M.P., Lansdorp-Vogelaar, I., Goede, S.L., Kuipers, E.J., Dekker, E., Stoker, J., van Ballegooijen, M., 2018. Colorectal cancer: cost-effectiveness of colonoscopy versus ct colonography screening with participation rates and costs. Radiology 287, 901–911.

31. Michalopoulos, N., Theodoropoulos, G., Stamopoulos, P., Sergentanis, T., Memos, N., Tsamis, D., Flessas, I., Menenakos, E., Kontodimopoulos, N., Zografos, G., 2013. A cost-utility analysis of laparoscopic vs open colectomy of colorectal cancer in a public hospital of the greek national health system. J BUON 18, 86–97.

32. Murphy, J., Halloran, S., Gray, A., 2017. Cost-effectiveness of the faecal immunochemical test at a range of positivity thresholds compared with the guaiac faecal occult blood test in the nhs bowel cancer screening programme in england. BMJ open 7, e017186.

33. ÓCéilleachair, A., Costello, L., Finn, C., Timmons, A., Fitzpatrick, P., Kapur, K., Staines, A., Sharp, L., 2012. Inter-relationships between the economic and emotional consequences of colorectal cancer for patients and their families: a qualitative study. BMC gastroenterology 12, 62.

34. ÓCéilleachair, A., Hanly, P., Skally, M., O’Leary, E., O’Neill, C., Fitzpatrick, P., Kapur, K., Staines, A., Sharp, L., 2017. Counting the cost of cancer: out-of-pocket payments made by colorectal cancer survivors. Supportive Care in Cancer 25, 2733–2741.

35. Pettersson, K., Carlsson, G., Holmberg, C., Sporrong, S.K., 2012. Cost identification of nordic fliri, nordic flox, xeliri and xelox in first-line treatment of advanced colorectal cancer in sweden–a clinical practice model approach. Acta Oncologica 51, 840–848.

36. Pil, L., Fobelets, M., Putman, K., Trybou, J., Annemans, L., 2016. Cost-effectiveness and budget impact analysis of a population-based screening program for colorectal cancer. European journal of internal medicine 32, 72–78.

37. Pilgrim, H., Tappenden, P., Chilcott, J., Bending, M., Trueman, P., Shorthouse, A., Tappenden, J., 2009. The costs and benefits of bowel cancer service developments using discrete event simulation. Journal of the Operational Research Society 60, 1305–1314.

38. PRISMA, 2020. Preferred reporting items for systematic reviews and meta-analyses (prisma). http://prisma-statement.org/ Accessed: 20.07.2020.

39. Rao, C., Smith, F., Martin, A., Dhadda, A., Stewart, A., Gollins, S., Collins, B., Athanasiou, T., Myint, A.S., 2018. A cost-effectiveness analysis of contact x-ray brachytherapy for the treatment of patients with rectal cancer following a partial response to chemoradiotherapy. Clinical Oncology 30, 166–177.

40. Rautenberg, T., Siebert, U., Arnold, D., Bennouna, J., Kubicka, S., Walzer, S., Ngoh, C., 2014. Economic outcomes of sequences which include monoclonal antibodies against vascular endothelial growth factor and/or epidermal growth factor receptor for the treatment of unresectable metastatic colorectal cancer. Journal of medical economics 17, 99–110.

41. Robles-Zurita, J., Boyd, K.A., Briggs, A.H., Iveson, T., Kerr, R.S., Saunders, M.P., Cassidy, J., Hollander, N.H., Tabernero, J., Segelov, E., et al., 2018. Scot: a comparison of costeffectiveness from a large randomised phase iii trial of two durations of adjuvant oxaliplatin combination chemotherapy for colorectal cancer. British journal of cancer 119, 1332.

42. Sobhani, I., Alzahouri, K., Ghout, I., Charles, D.J., Durand-Zaleski, I., 2011. Cost-effectiveness of mass screening for colorectal cancer: choice of fecal occult blood test and screening strategy. Diseases of the Colon & Rectum 54, 876–886.

43. Špacírová, Z., Epstein, D., García-Mochón, L., Rovira, J., de Labry Lima, A.O., Espín, J., 2020. A general framework for classifying costing methods for economic evaluation of health care. The European Journal of Health Economics, 1–14.

44. Tilson, L., Sharp, L., Usher, C., Walsh, C., Whyte, S., O’Ceilleachair, A., Stuart, C., Mehigan, B., Kennedy, M.J., Tappenden, P., et al., 2012. Cost of care for colorectal cancer in ireland: a health care payer perspective. The European journal of health economics 13, 511–524.

45. Tscheulin, D.K., Drevs, F., 2010. The relevance of unrelated costs internal and external to the healthcare sector to the outcome of a cost-comparison analysis of secondary prevention: the case of general colorectal cancer screening in the german population. The European Journal of Health Economics 11, 141–150.

46. Whyte, S., Chilcott, J., Halloran, S., 2012. Reappraisal of the options for colorectal cancer screening in england. Colorectal Disease 14, e547–e561.

47. Woollard, M., 2014. 3.1 administrative data: Problems and benefits. a perspective from the united kingdom1. Facing the Future: European Research Infrastructures for the Humanities and Social Sciences, 49.

48. York Health Economics Consortium, 2016. A glossary of health economic terms. https://yhec.co.uk/resources/glossary/ Accessed: 1.06.2020.

